# Explainable artificial intelligence for cough-related quality of life impairment prediction in asthmatic patients

**DOI:** 10.1101/2023.10.04.23296540

**Authors:** Sara Narteni, Ilaria Baiardini, Fulvio Braido, Maurizio Mongelli

## Abstract

Explainable Artificial Intelligence (XAI) is becoming a disruptive trend in healthcare, allowing for transparency and interpretability of autonomous decision-making. In this study, we present an innovative application of a rule-based classification model to identify the main causes of chronic cough-related quality of life (QoL) impairment in a cohort of asthmatic patients. The proposed approach first involves the design of a suitable symptoms questionnaire and the subsequent analyses via XAI. Specifically, feature ranking, derived from statistically validated decision rules, helped in automatically identifying the main factors influencing an impaired QoL: pharynx/larynx and upper airways when asthma is under control, and asthma itself and digestive trait when asthma is not controlled. Moreover, the obtained *if-then* rules identified specific thresholds on the symptoms associated to the impaired QoL. These results, by finding priorities among symptoms, may prove helpful in supporting physicians in the choice of the most adequate diagnostic/therapeutic plan.

## Introduction

Nowadays Artificial Intelligence (AI) is revolutionizing medicine by leveraging powerful technologies and advanced learning algorithms. This has the potential to support several clinical processes, from prognostics to diagnostics, from treatment management to drug discovery, and also can aid hospital administrative tasks. However, AI real application in healthcare needs to be approached very carefully, since failures may cause harm to human lives. For this reason, AI research is increasing its interests in *trustworthy AI* [1], a broad paradigm establishing how to properly design, develop and deploy real-world AI applications. Between its principles, *transparency* requires providing the user with an understanding of the autonomous decisions generated by the model: this topic is subject of *eXplainable AI (XAI)* research [2, 3]. XAI comprehends a wide range of methodologies, which can be broadly categorized as post-hoc explanations of black box models and transparent-by-design techniques [4]. In the latter category, rule-based models are characterized by understandable decision rules expressed in the *if-then* format. These kinds of models are particularly suitable in medicine, since their intrinsic interpretability allows clinicians to enter models’ logic and increase trust in them. In light of this, our work focuses on the usage of such techniques to characterize the quality of life of asthmatic patients with chronic cough.

Asthma is a frequent cause of cough in adults [5]. In addition to coughing, asthmatic patients may also wheeze or feel short of breath. However, some people have a condition known as cough variant asthma, in which cough is the only symptom of asthma. For these reasons, tools for the assessment of asthma, such as Asthma Control Test (ACT) [19], consider cough among the asthma features. While in patients with uncontrolled asthma the disease itself can be the cause of cough, the persistence of cough despite good asthma control can be related to concomitant disorders (i.e., postnasal drip, pharynx/larynx disorders, and acid reflux from the stomach [6]) or inability of asthma drugs to fully remove the symptoms.

In light of these considerations, it is very useful to design a method that allows to define the priority of choice among different diagnostic techniques, starting from patients’ self-reported presence and entity of symptoms and their impact on the quality of life. Methods based on XAI, thanks to their transparent and interpretable methods, can offer a great opportunity in this direction.

### Contribution

In this study, we propose the usage of a rule-based XAI model to support clinicians in the diagnostic procedure for determining the origins of chronic cough in asthmatic patients. More precisely, our main contributions are the following:

- We introduce a new block-based questionnaire, devoted to collect (respiratory) symptoms perceived by asthmatic patients with chronic cough.
- We train a rule-based model, the Logic Learning Machine (LLM), for predicting chronic cough-related quality of life based only on self-reported responses to the questionnaire of symptoms, by distinguishing patients with high or low asthma control level.
- By validating and analyzing the model, we discover which symptoms and corresponding values are mainly involved in a quality of life exacerbation.

The remaining part of the paper is organized as follows. In Section Related Work we report some recent examples of machine learning for chronic cough. Section Methodology describes the workflow, the dataset structure and the adopted methodologies. Section Results shows and discusses the obtained results. Finally, Section Conclusion concludes the paper and reports future research on the topic.

## Related Work

Different machine learning (ML) and AI-based studies on chronic cough and asthma have been carried out in recent years, by leveraging the newest medical technologies [17]. An AI-based cough count, CoughyTM [14], system was recently developed that quantifies cough sounds collected through a smartphone application. Study results showed that suggest that CoughyTM could be a novel solution for objectively monitoring cough in a clinical setting. A vocal biomarker-based machine learning approaches have shown promising results in the detection of various health conditions, including respiratory diseases, such as asthma [15]. Also, a deep learning model for identifying chronic cough patients with even higher sensitivity and specificity when structured and unstructured electronic health records EHR data are utilized has been proposed [16].

In [12], well established ML models like gradient boost and random forest were adopted in a retrospective study to predict the risk of persistent chronic cough (PCC) in patients with chronic cough (CC). The work proposed in [7] used a statistical approach (Latent Class Analysis) on the Swedish Twin study On Prediction and Prevention of Asthma (STOPPA) and the Child and Adolescent Twin Study in Sweden (CATSS) questionnaires responses to identify asthma and wheeze phenotypes in children. In [9], four adult chronic cough phenotypes were identified through a cluster analysis method applied to questionnaire data such as the COugh Assessment Test (COAT) [10] and the Korean version of the Leicester Cough Questionnaire [11].

However, all these literature examples do not provide their outcomes in an explainable way.

## Methodology

### Workflow

The overall methodology followed in the proposed analyses is depicted in Fig. 1. The dataset was first split in a 70% training and 30% test sets, then an explainable Artificial Intelligence (XAI) model was considered for data classification. The adopted classifier is called Logic Learning Machine and provides its predictions through a set of rules. In order to verify the statistical significance of the resulting ruleset, this was validated through a statistical test. Rules that did not pass the test were then filtered out from the model, thus obtaining a final, validated, set of rules. Also, feature ranking was investigated to identify which of the inputs have the higher impact on the model outcome. Finally, the overall performance of the validated ruleset was measured on the test set, by considering some common metrics for machine learning models evaluation.

**Fig 1.**
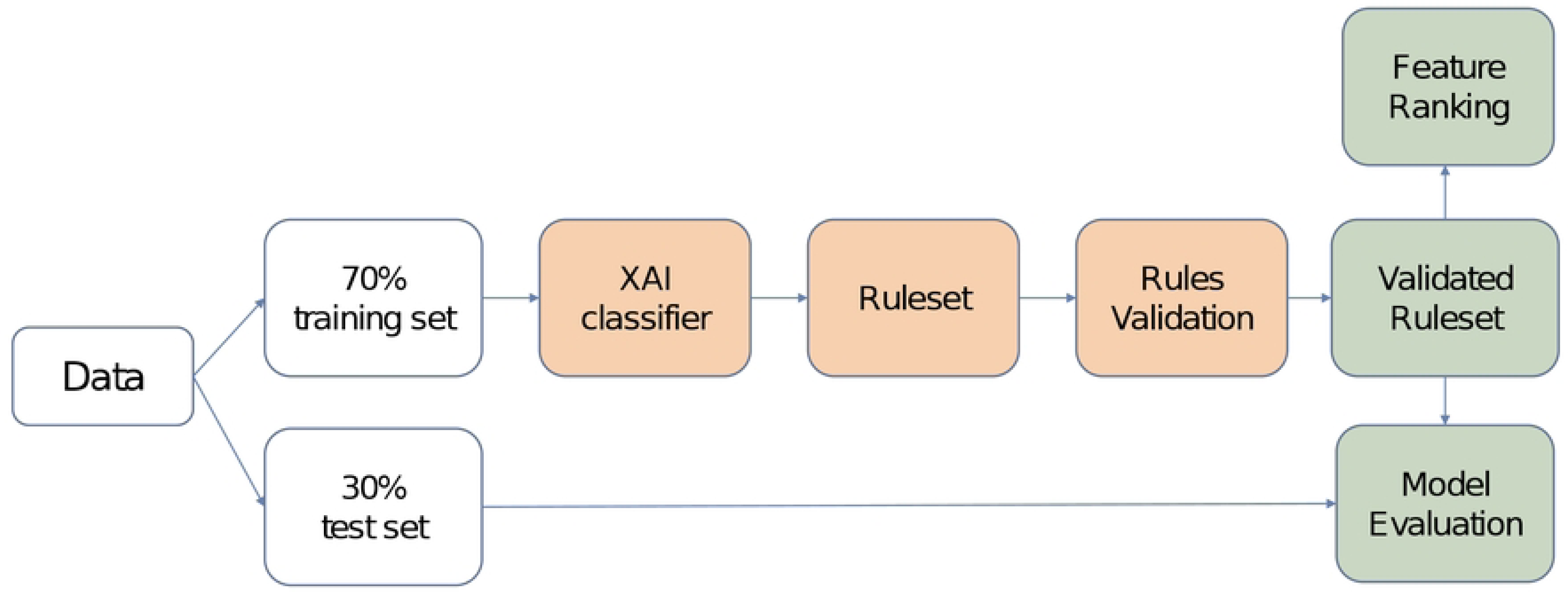
Workflow of the analyses carried out in the proposed XAI-based approach.

Next Sections provide the description of the dataset and some fundamentals about the adopted XAI, the rule validation test and the definition of the evaluation metrics.

### Dataset Description

The study involved a cohort of asthmatic patients, who have been asked to answer to three different kinds of questionnaires (data were accessed on 2023/03/08; the authors had no access to information that could allow to identify individual participants during or after data collection).

The first one collects patients’ feedback about a variety of symptoms. Specifically, it contains 19 items relating to four domains related to the more frequent causes of chronic cough, as shown in the diagram of Fig. 2.

**Fig 2.**
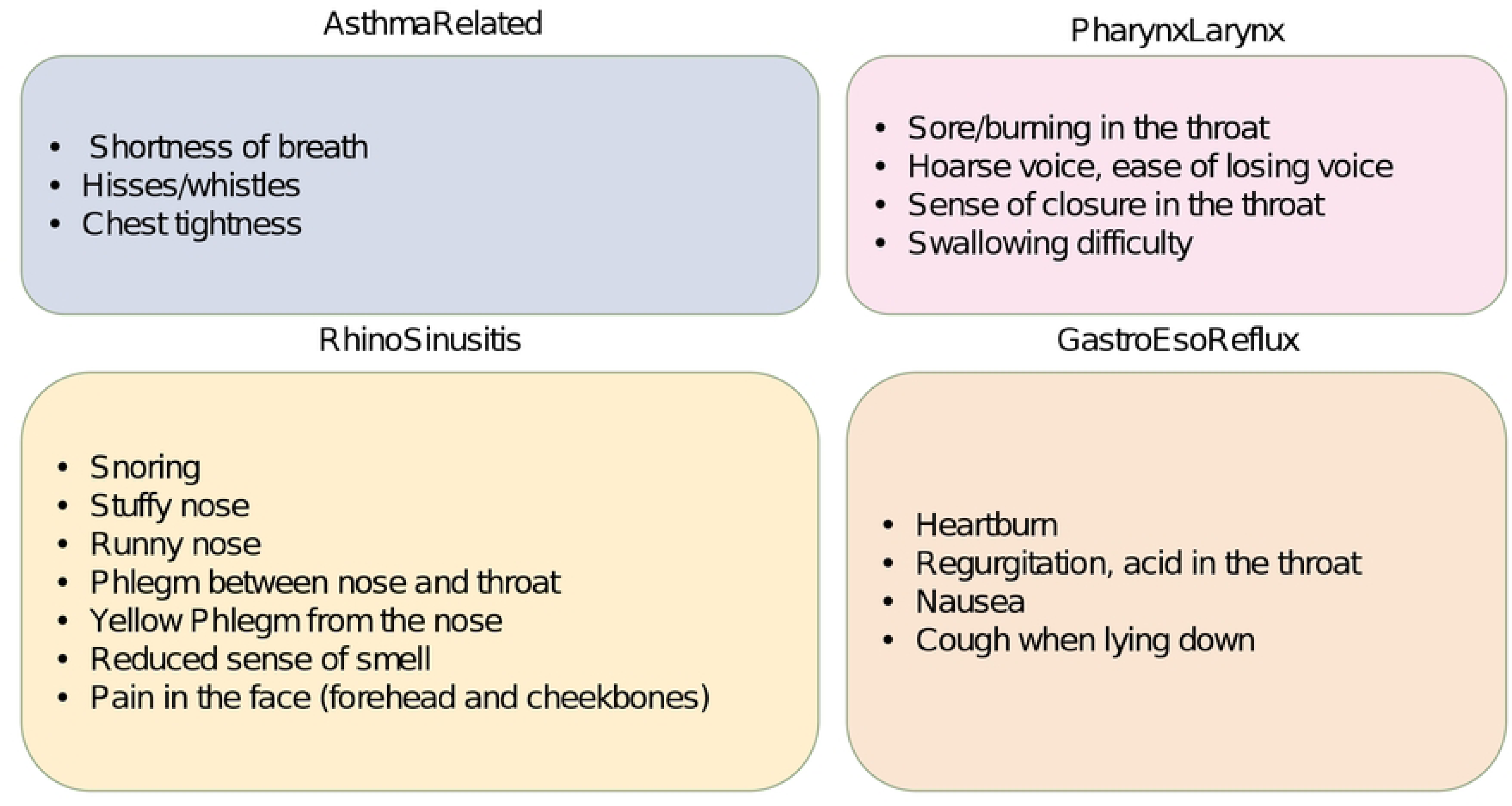
Symptoms questionnaire. Schematic representation of the four blocks (*AsthmaRelated, PharynxLarynx, RhinoSinusitis, GastroEsoReflux*) of the symptoms questionnaire and their related items.

For each item, the patients answered to the question *“How intense/annoying has the symptom been in the last month?”*, by self-reporting a level between *None* and *Very Much* expressing the perceived entity of the corresponding symptom. These levels were then proportionally converted to a score in the 0-100 scale. The average of the responses within each block was computed, thus individuating a set of four features that will be used as input to the ML model, each referred to a different body organ.

The second questionnaire involved in this study is the Chronic Cough Impact Questionnaire (CCIQ) [18]. It is useful to measure the impact of cough on health-related quality of life, namely *impact on daily life (CCIQ IDL)*, on *sleep/concentration (CCIQ SC)*, on *mood (CCIQ M)* and *relationship (CCIQ R)*. A score for each group is derived and contributes to compute a global score, called *CCIQ GLS* : based on this, we defined two classes of patients. Those scoring *CCIQ GLS* ≥ 20 were labelled as *impaired Quality of Life (QoL)*, while those with *CCIQ GLS <*20 were associated to a *near normal QoL*. The last questionnaire considered is the Asthma Control Test (ACT) [19]. It is a item questionnaire aimed at assessing at which extent the asthmatic patient has control of the pathology. We used the score obtained from this test to further distinguish patients between two populations: subjects with *ACT* ≥ 20 were identified as the *controlled asthma* group, whereas those scoring *ACT <* 20 formed the *not controlled asthma* group.

The analyses carried out in this work thus considered three different cases: i) *all* patients were included; ii) only *controlled* asthma patients were included; iii) only *not controlled* asthma patients were included.

### The Adopted eXplainable AI classifier

For each patients group, we trained a XAI classifier that, fed with the 4 input features (referred to as *AsthmaRelated, PharynxLarynx, RhinoSinusitis* and *GastroEsoReflux*) representing the average scores on each block of the symptoms questionnaire (Fig. 2), provided a prediction of the patient’s cough-related QoL, which can be either *impaired* or *near normal*.

The analyses on the first group (i.e., *all* patients) did not explicitly use the knowledge acquired from the ACT questionnaire. Indeed, the classification model that is designed for this group represents a tool to individuate which areas and values of symptoms drive an impaired QoL in a generic asthmatic population, but without any previous knowledge on the asthma control level. Conversely, the analyses performed on the *controlled asthma* and *not controlled asthma* groups also exploited the information from the ACT, thus the results of the XAI predictive models provide indications that are specifically tailored to the different asthma control level.

#### Logic Learning Machine

In this Section, we provide some basic description of the adopted classifier, the Logic Learning Machine (LLM). It is a rule-based explainable AI model, designed and developed by Rulex [26] as the efficient implementation of Switching Neural Networks [20].

Given the input data, the LLM provides a classification model described by a set of rules 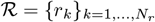, where each *r*_*k*_ is expressed with the form: **if** *<premise>* **then** *<consequence>*. The *<premise>* constitutes the antecedent of the rule and is a logical conjunction (AND) of conditions on the input features. The *<consequence>* reports the outcome of the classification, i.e. the predicted class label.

The performance of any rule *r*_*k*_ ∈ ℛ can be evaluated by covering *C*(*r*_*k*_) and error *E*(*r*_*k*_) metrics, defined as:

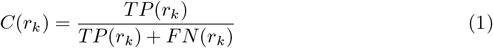

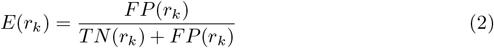

where *TP* (*r*_*k*_) and *FP* (*r*_*k*_) are the number of samples that, respectively, correctly and wrongly verify rule *r*_*k*_; *TN* (*r*_*k*_) and *FN* (*r*_*k*_) are the number of samples that, respectively, correctly and wrongly do *not* verify the rule. The covering is also proportional to the relevance of the rule, therefore the larger it is, the higher is the probability that the rule is valid on new unseen samples. On the contrary, the error *E*(*r*_*k*_) measures how much wrongly covered is the rule and its maximum value is usually fixed as a model hyperparameter (by default, it is of 5%).

Both covering and error are useful to define *feature ranking*. It allows to gain insights on which input attributes contribute the most to predict a given class; to this aim, values of relevance for each feature are computed and typically represented in bar plots in descending order.

Given a feature *X*_*j*_ and a rule *r*_*k*_ (predicting class label *ŷ*) containing in its premise a condition *c*_*j*_ on variable *X*_*j*_, covering and error are first combined to compute the relevance of *c*_*j*_ as 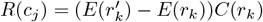, where 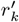 is the rule obtained by removing condition *c*_*j*_ from *r*_*k*_. The relevance 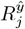 for feature *X*_*j*_ is then derived by the following equation 3:

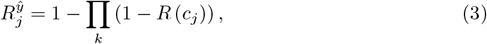

where the product is computed on the rules *r*_*k*_ that include a condition *c*_*j*_ on the feature of interest.

### Rules Statistical Validation

In order to assess the statistical significance of the set of rules generated by the LLM, we decided to use the Pearson’s *χ*^2^ independence test [23]. To this purpose, we considered two binary events involving the available data samples, namely their membership to an output class and their satisfaction of the rules in ℛ. Therefore, a 2×2 contingency table was built for each rule *r*_*k*_ ∈ ℛ, as shown in Table 1, reporting the counts of how many samples of the two classes are covered or not by the rule.

**Table 1.** 2×2 contingency matrix for rule *r*_*k*_.

| | $\vdash r_k$ | $\not\vdash r_k$ |
| --- | --- | --- |
| $y = 1$ | $a$ | $b$ |
| $y = 0$ | $c$ | $d$ |

Let the input dataset be 𝒯 = {(**x**_*i*_, *y*_*i*_)}_*i*=1,…,*N*_, with binary output labels *y*_*i*_ = 0 (i.e., *near normal QoL* class in our case) or *y*_*i*_ = 1 (i.e., *impaired QoL* class). Also, let us define with **x**_*i*_ ⊢ *r*_*k*_ and **x**_*i*_ ⊬ *r*_*k*_ the satisfaction and unsatisfaction of rule *r*_*k*_ by the data point **x**_*i*_, respectively. Then, the following quantities can be defined:

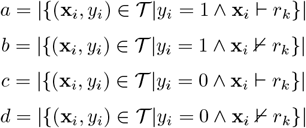

*χ*^2^ statistic was then computed starting from the matrix. The test was carried out with a null hypothesis of independence between class label and rule membership, with a significance level of 0.05 for the *p*-value. Rules with a *p*-value *<* 0.05 were then proved as statistically significant [24] and those that did not pass the test were removed from the ruleset ℛ, giving rise to a set of validated rules ℛ_*val*_ ⊆ ℛ.

### Model Performance Evaluation

To evaluate the overall performance of the validated ruleset, the confusion matrix reporting the True Positives (TP, i.e., patients correctly predicted as *impaired QoL*), False Positives (FP, i.e., *near normal QoL* patients wrongly predicted as *impaired QoL*), True Negatives (TN, i.e., patients correctly predicted as *near normal QoL*) and False Negatives (FN, i.e., *impaired QoL* patients wrongly predicted as *near normal QoL*) obtained by applying such rules to a test set was first built. It is the basis to define the following measurements, particularly useful when evaluating the outcomes of a clinical ML model [25]:

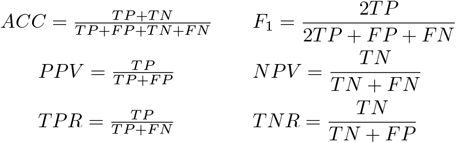

While accuracy (ACC) and *F*_1_-score (F_1_) provide an evaluation of the model taking into account its performance on both the classes, the other ones assess the performance on single classes. In detail, Positive Predictive Value (or precision, PPV) and True Positive Rate (or sensitivity or recall, TPR) reflect the number of TPs over the total amount of positive predictions and the total amount of positive samples, respectively. Viceversa, Negative Predictive Value (NPV) and True Negative Rate (or specificity, TNR) represent the number of TNs over the total amount of negative predictions and the total quantity of negative samples, respectively.

## Results

This study involved a population of 283 asthmatic patients (i.e., the *all* group), with age 33.5±7.77 and characterized by a Forced Expiratory Volume in the first second (FEV1) of 96.5% ± 19.09 and an ACT score of 19.09±4.98. 146 patients belong to the *controlled asthma* group (i.e., the 52% of the whole population), while the remaining 137 patients form the *not controlled asthma* group.

### Data statistics at a first glance

Figure 3 provides a first glance on how the four blocks of symptoms are distributed between the two classes (impaired QoL and near normal QoL) both in the controlled and not controlled asthma patients. Each colored bar individuates a different group of patients and its length (the interquartile range, or IQR) varies between the 25th and 75th percentiles, while the vertical dashed lines (i.e., the whiskers) range from the minimum to the maximum values and, finally, the horizontal dot-dashed black line points out the median value of the corresponding symptoms group. The red ‘+’ markers represent outlier points. It is possible to observe that *PharynxLarynx* can better distinguish the two classes in the controlled asthma group, since the median value of one class falls outside the bar of the other. A similar reasoning holds for the not controlled asthma group, where *AsthmaRelated, PharynxLarynx* and *GastroEsoReflux* stand out. However, this kind of evaluation is based on visual analytics and simple statistics, and the results do not provide any guarantee of validity on new, unseen, patients. This is why we decided to rely on machine learning-based approaches.

**Fig 3.**
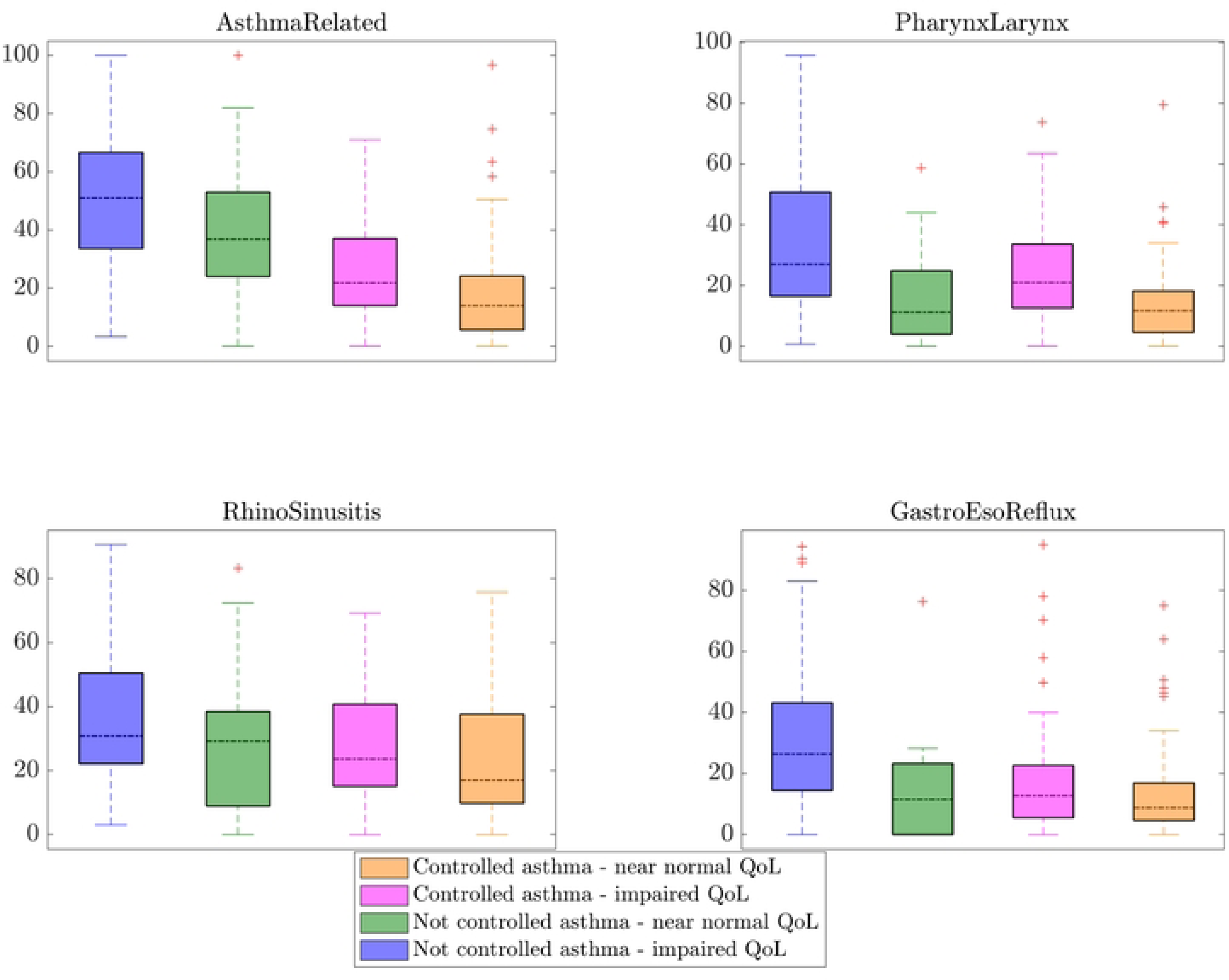
Box plots. Graphs showing the class distributions in the controlled versus not controlled patients groups, for each of the considered features.

### Explainable AI-based analysis

For each of the considered cases, the LLM algorithm was trained on a 70% training set and generated a set of rules. In particular, for the *all* group, 19 rules were generated (8 predicting *impaired QoL* class and 11 the *near normal QoL*); from the *controlled asthma* case, we got 13 rules (4 for the *impaired QoL* and 9 for the *near normal QoL* class); lastly, 9 rules derived from the *not controlled asthma* group (5 referring to the *impaired QoL* and 4 to the *near normal QoL* class).

The Pearson’s *χ*^2^ validation test was then carried out to statistically proof the obtained rulesets, as per the procedure detailed in Section. After the test, 2 rules out of 8 for the *impaired QoL* class and 4 out of 11 for the *near normal QoL* class were validated in the *all* case; 2 of the 4 rules predicting the *impaired QoL* class in the *controlled asthma* group resulted significant, while 3 out of 9 rules for the other class was validated in the same group; similarly, in the *not controlled asthma* patients, 2 rules out of 5 for the *impaired QoL* class passed the test, while 3 out of 4 rules related to the *near normal QoL* did.

#### Model performance metrics

After validating the rules, we thus have been able to define a final set of rules for each case, by leaving out from the original rulesets all those which tested not significant. The predictive performance of the validated rulesets was assessed on the test set, by computing the metrics described in Sec. ; their values are depicted and compared in Fig. 4 for the three groups.

**Fig 4.**
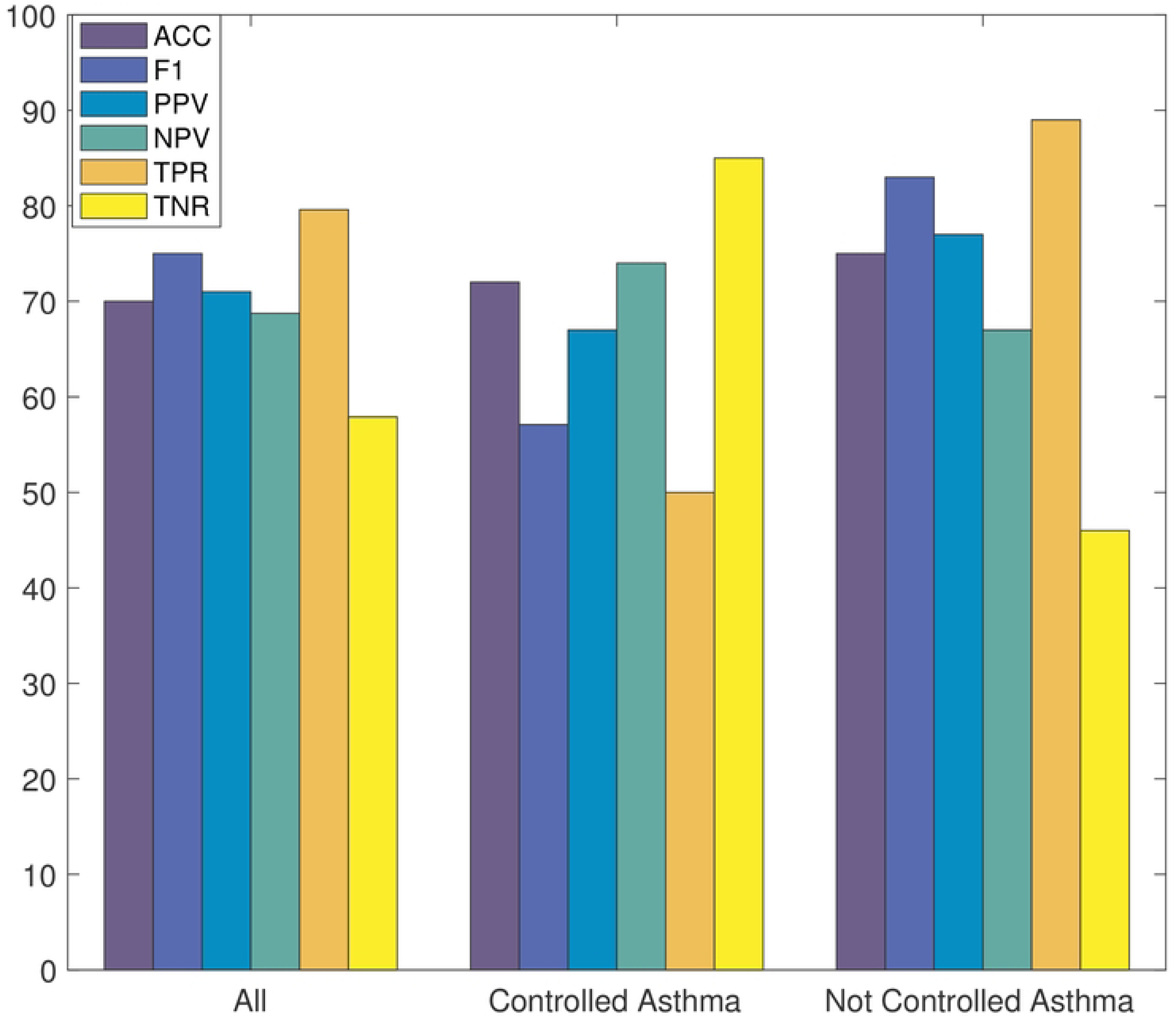
Validated rules performance. Percentage values of the accuracy (ACC), F_1_-score (F1), Positive Predictive Value (PPV), Negative Predictive Value (NPV), True Positive Rate (TPR) and True Negative Rate (TNR) of the LLM in the three patients’ groups.

The accuracy reached at least 70% in all cases, thus showing good performance of the validated rulesets. While also F_1_ score value was high for the *all* and *not controlled asthma* groups (75% and 83%, respectively), it was lower (57%) for the *controlled asthma* group, denoting both poorer precision and recall. Indeed, PPV and TPR metrics, related to the positive class (i.e., impaired QoL), were found 66% and 50%, respectively, whereas NPV and TNR (reflecting the model’s performance on the negative class, i.e., the near normal QoL) were sensitively larger (74% and 85%, respectively). In contrast, the *not controlled asthma* reached a high F_1_ due to larger values of precision and recall, with a PPV of 77% and TPR of 89%; on the other hand, NPV and TNR resulted in lower values. A similar reasoning holds for the *all* group, even if the model performance on the two classes was more balanced, with less difference among the metrics for the positive and the negative class.

#### Most relevant symptoms questionnaire items

Further insights on the LLM results were obtained by visualizing the feature ranking. Bar plots, obtained for the three cases under analysis, are shown in Fig. 5, representing the *impaired QoL* class feature ranking, that highlights which of the features influenced more the LLM decision towards that class. Concerning the *all* group, from Fig. 5A *AsthmaRelated* and *PharynxLarynx* were individuated as the main factors leading to an impaired cough-related quality of life. In contrast, the main attributes for the *controlled asthma* group (Fig.5B) were *PharynxLarynx* and *RhinoSinusitis*. Finally, dominant features for the *not controlled asthma* resulted *AsthmaRelated* and *GastroEsoReflux* (Fig. 5C).

**Fig 5.**
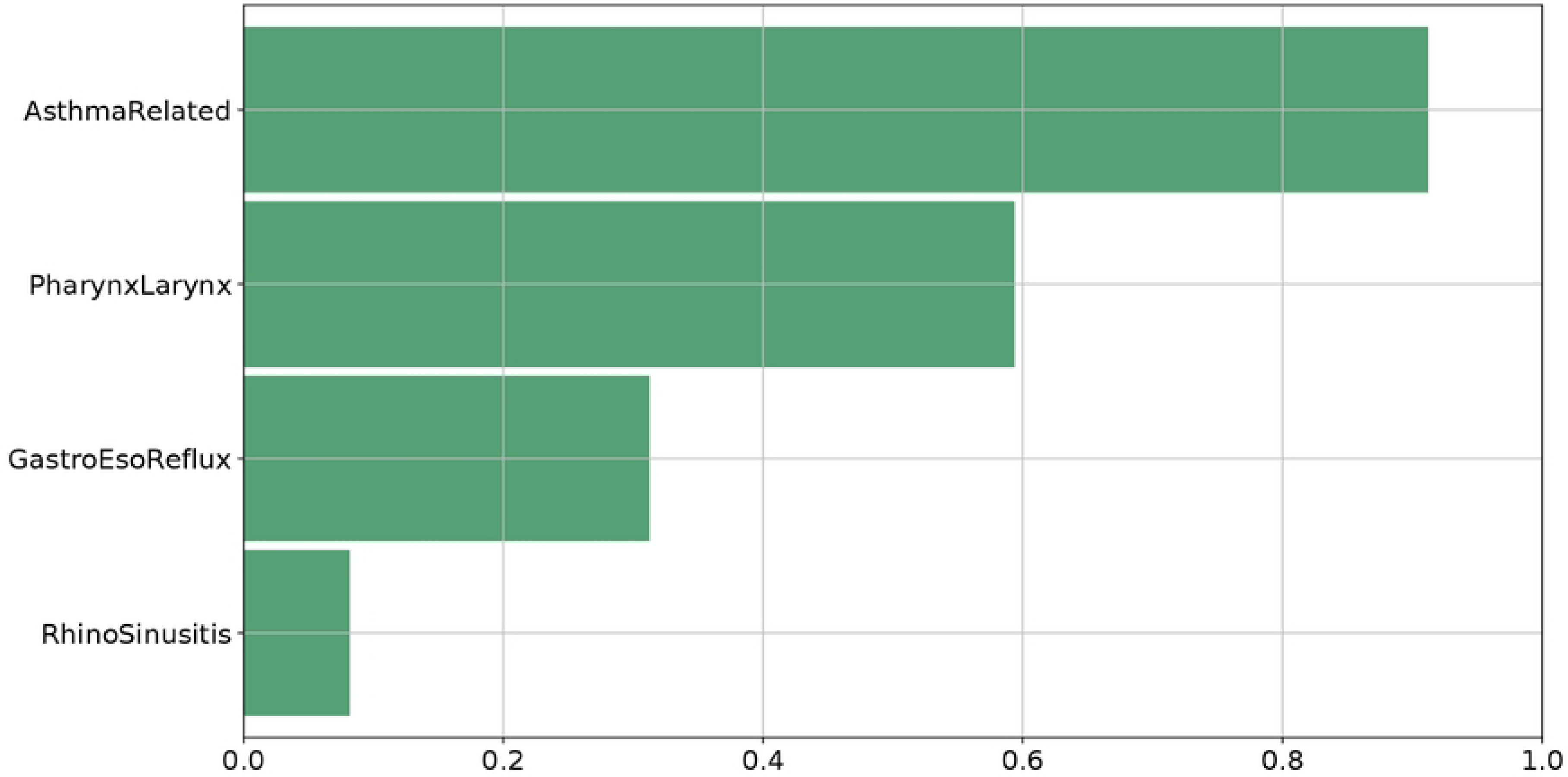
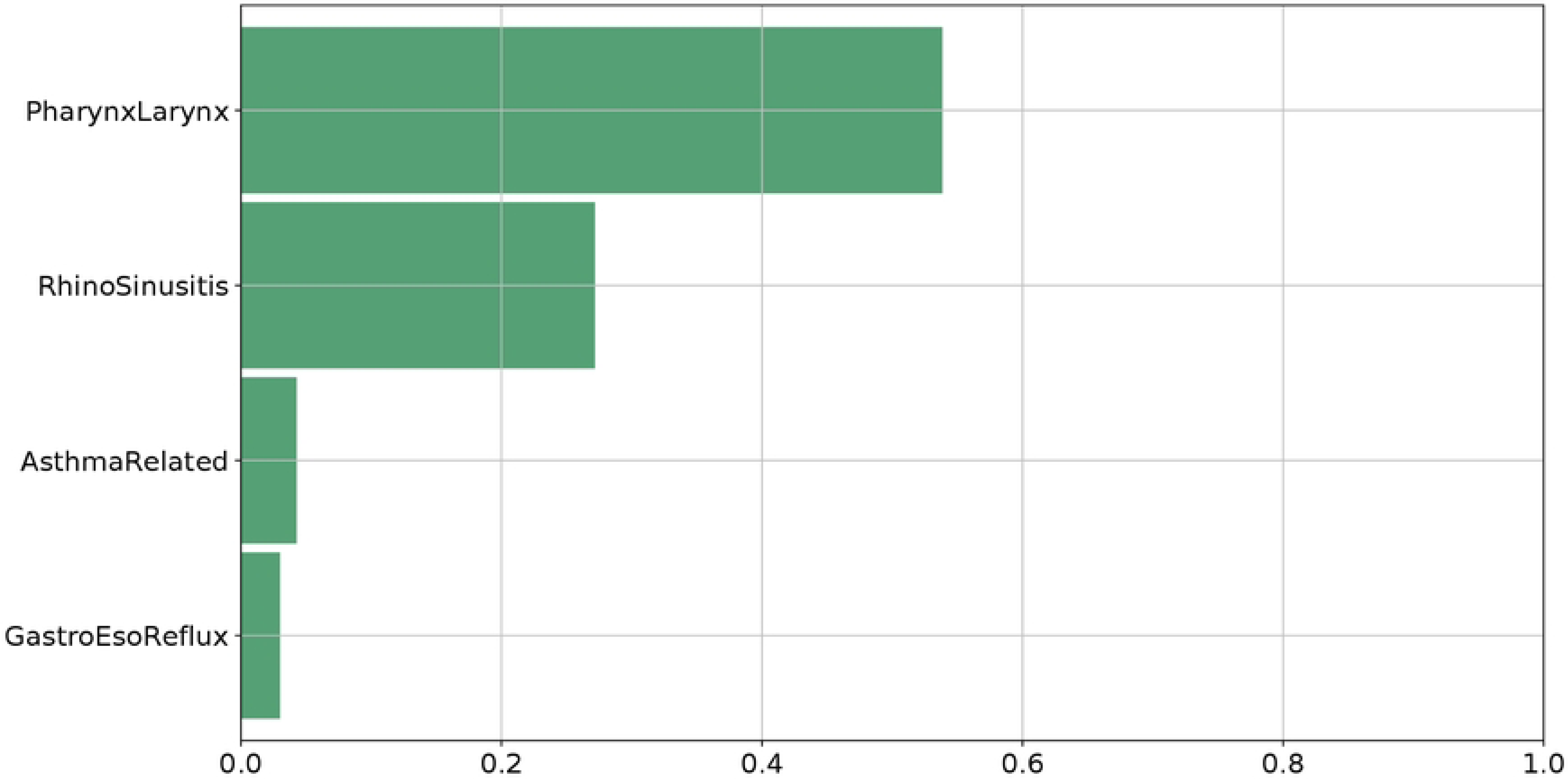
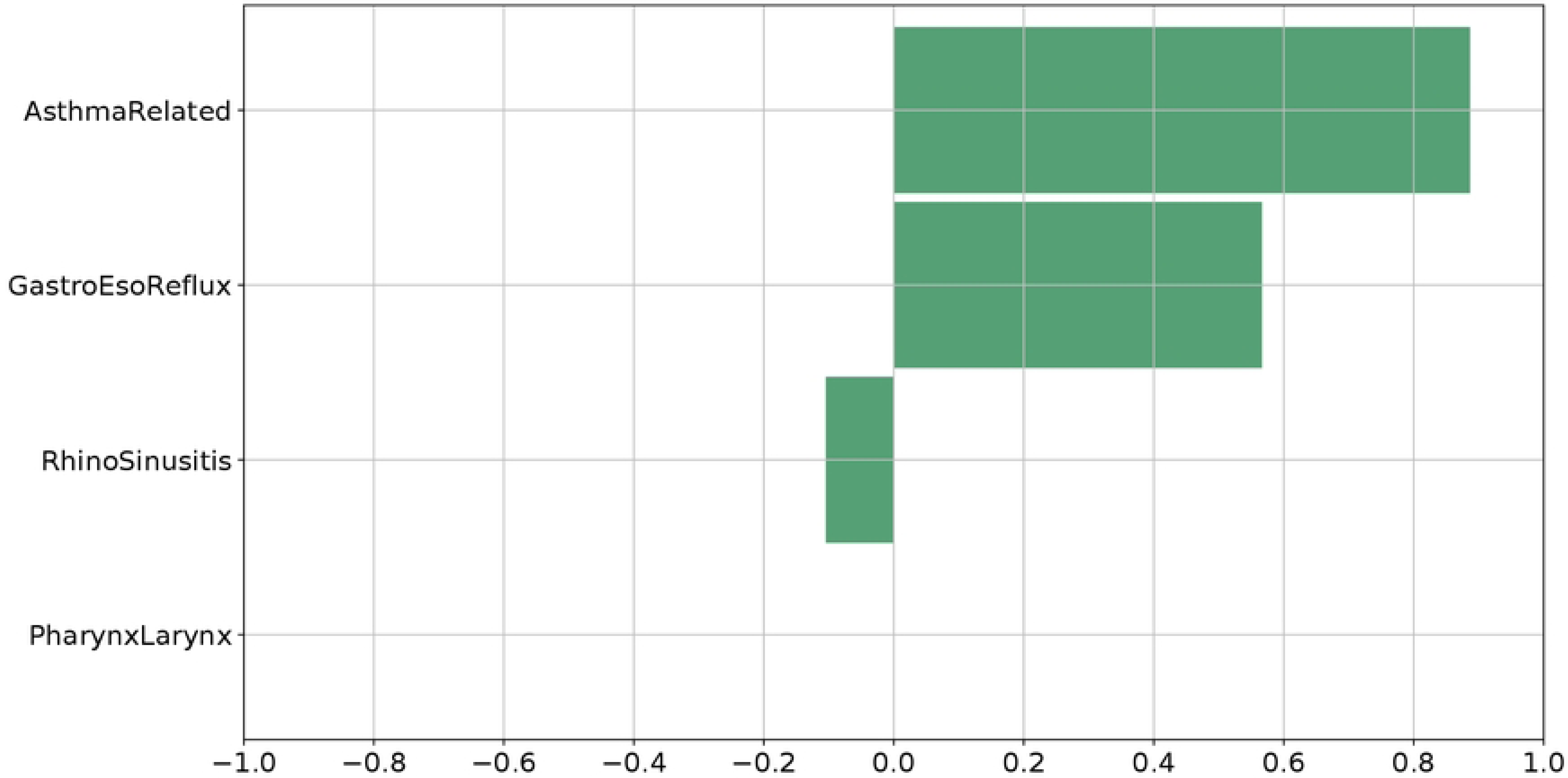
Feature Ranking. LLM feature ranking for the impaired QoL class in the three cases. (A): all group; (B): controlled asthma group; (C): Not controlled asthma group.

The presence of *AsthmaRelated* as a relevant factor for the *not controlled asthma* group is in line with our expectation, since the deterioration of these patients’ QoL reasonably depends on the asthma itself and the clinical investigation should be primarily addressed to it. Secondarily, the digestive tract should be considered. Conversely, the feature ranking for the *controlled asthma* patients provides the indication that further clinical assessments should focus first on the throat and, then, on the nose. By using the symptoms questionnaire, in absence of any information about the patient’s asthma control level, results suggest to first consider the asthma and then the nose.

#### Symptoms questionnaire scores driving impaired QoL

While previous Section provided which are the main factors involved in the impaired QoL, in this Section our focus is posed on the information we can derive by inspecting the validated rules predicting the *impaired QoL* class, which are reported in Table 2. Their aim is to define useful criteria to support clinicians in the diagnostic process, by individuating, in the three cases, which values assumed by the symptoms questionnaire scores are more probably associated to an *impaired QoL* status. However, by looking at the threshold values of a same indicator in the two rules for a given group, it can be noticed that they can be pretty different or even conflicting. For example, the 8.16 and the 28.16 in the *AsthmaRelated* score for the *all* group have a difference of 20 percentage points, which cannot be disregarded; also, the condition on *PharynxLarynx* in the *controlled asthma* group is discordant in the two related rules, the first stating that values larger than 23.46 lead to QoL deterioration, while the second states the same for values lower than 13.25. Regarding the *not controlled asthma* case, the two rules seem to individuate two clusters of patients, one depending on increasing (*>* 50.83) *AsthmaRelated* score and decreasing (≤70.43) *RhinoSinusitis* score, and the other depending on *GastroEsoReflux* score only. Therefore, rule generation alone is able to individuate several clusters of patients, each described by a pretty different set of conditions on the questionnaire scores. Nevertheless, our final goal is to provide, through the ML system, more general information to be used in clinical practice, especially valid in the case of new, never seen before, patients.

**Table 2.** Criteria for *impaired QoL* prediction through symptoms questionnaire, as emerged from LLM rules validated through the *χ*^2^ independence test, for each considered patient group. Pink-colored cells highlight the rules that were proved the most performing even on previously unseen patients.

| Case | Significant Rules | Covering (%) | Error (%) |
| --- | --- | --- | --- |
| All | 1. **if** *AsthmaRelated* > 8.16 **and** *PharynxLarynx* > 15.12 **and** *RhinoSinusitis* ≤ 70.43 **and** *GastroEsoReflux* > 8.62 **then** *impaired QoL* | 57 | 4.1 |
|  | 2. **if** *AsthmaRelated* > 28.16 **and** 7.93 < *RhinoSinusitis* ≤ 70.43 **and** *GastroEsoReflux* > 4.62 **then** *impaired QoL* | 56 | 4.1 |
| Controlled Asthma | 1. **if** *AsthmaRelated* > 10.83 **and** *PharynxLarynx* > 23.46 **and** *RhinoSinusitis* ≤ 69.81 **then** *impaired QoL* | 47 | 3.7 |
|  | 2. **if** *PharynxLarynx* ≤ 13.25 **and** 11.57 < *RhinoSinusitis* ≤ 26.78 **and** *GastroEsoReflux* > 0.62 **then** *impaired QoL* | 21 | 3.7 |
| Not controlled Asthma | 1. **if** *AsthmaRelated* > 50.83 **and** *RhinoSinusitis* ≤ 70.43 **then** *impaired QoL* | 52 | 0.0 |
|  | 2. **if** *GastroEsoReflux* > 28.375 **then** *impaired QoL* | 41 | 0.0 |

Further evaluations of the models are then carried out for a better knowledge extraction suitable to our objective. Covering and error percentages reported in the Table have been derived during model training on the training data portion. Hence, their values, even when considerably high (as in the cases of *>* 50% covering), do not guarantee the same performance on test (previously unseen) data. Thus, percentages of impaired QoL test points satisfying either one, both or even none of the two rules were computed to understand how the original covering changes on new data; the obtained values are outlined in Table 3. When points satisfy both rules, the most important one can still be individuated as the highest-covering one (from Tab. 2). Thus, the 44.90% rate of satisfaction of both rules in the *all* group contributes to the rate of rule 1 (of the same group), which then reaches a total value of 65.31% of satisfaction. Thus, this rule should be taken as a reference for individuating the factors with higher impact on the impaired QoL. The same reasoning holds for the *not controlled asthma* group, where rule 1 reaches the about the 52%. Regarding the *controlled asthma* case, rule 1 proves as the most frequently validated by the unseen patients. Moreover, it is worth noting that the sum of the percentages shown in Table for Rule 1, Rule 2 and Both rules columns corresponds to the TPR computed in Fig. 4. Hence, in this analysis we can see the specific contribution of the two rules in determining its value.

**Table 3.** Satisfaction percentages of validated rules for the *impaired QoL* class on unseen data. For each group, Rule 1 refers to rule number 1 of Table 2, and, similarly, Rule 2 here refers to rule number 2.

|  | **Rule 1** | **Rule 2** | **Both rules** | **No rules** |
| --- | --- | --- | --- | --- |
| **All** | 20.41 % | 14.28% | 44.90% | 20.41% |
| **Controlled asthma** | 41.67% | 8.33% | 0% | 50% |
| **Not controlled asthma** | 37.04% | 37.04% | 14.81% | 11.11% |

In summary, for each of the three groups, a rule has emerged as the one with the best predictive ability for an *impaired QoL* status and it can be considered as a helpful decision-making support for clinicians, especially at the beginning of the clinical evaluation process. Indeed, by using the information from the feature ranking (Fig. 5), we discovered the main blocks of symptoms associated to an impaired QoL status due to chronic cough and the individuated decision rules define which ranges of values should be considered alarming on those variables.

## Conclusion

In this work, we proposed the evaluation of the quality of life of asthmatic patients, with lower or higher degree of asthma control, experiencing chronic cough. To this end, we first developed a questionnaire to collect patients’ symptoms in relation to the most frequent causes of chronic cough (i.e., upper airways, pharynx/larynx, digestive tract, lower airways). The LLM-based analysis of patients’ responses to the questionnaire items, through feature ranking, helped in automatically identifying priorities among these causes: pharynx/larynx and upper airways when asthma is sufficiently controlled, and asthma itself and digestive trait when asthma is not controlled. Moreover, the adopted rule-based model, with proper statistical validation, identified which specific values of the symptoms are associated to an impairment of cough-related quality of life. The obtained results could support the physician in choosing the right diagnostic/therapeutic plan. However, sensitivity and specificity of the developed model need to be verified in further prospective studies. Furthermore, future research in this direction may investigate the adoption of other rule-based models than the LLM, as well as the usage of black-box algorithms with subsequent rule extraction.

## Data Availability

The dataset will be made available in a public Github repository

